# An independent, multi-timepoint evaluation of Disconnection Symptom Discoverer cognitive outcome prediction accuracy in stroke

**DOI:** 10.64898/2026.05.20.26353733

**Authors:** Lizzy Kenny, Margaret Jane Moore, Nele Demeyere

**Affiliations:** Nuffield Department of Clinical Neurosciences, University of Oxford, John Radcliffe Hospital, Oxford, UK; Queensland Brain Institute, University of Queensland, Brisbane, Australia

**Keywords:** stroke, outcome prediction, disconnection, post-stroke cognitive impairment, recovery

## Abstract

The Disconnection Symptom Discoverer (DSD) model proposes to predict long-term performance on neuropsychological tests from stroke lesion disconnection profiles. The model requires external validation to determine reproducibility and generalizability to new and different patients. Here, we investigated whether the DSD supports accurate multi-domain cognitive outcome predictions at three different timepoints post stroke, in a clinically representative independent cohort.

In this study, the DSD was used to predict visuospatial attention, verbal memory, and language scores in an independent cohort of 74 stroke survivors (mean age = 69.2, 39% female) with 3 repeated cognitive assessments. DSD-predicted scores were compared to observed neuropsychological scores collected at <2 weeks, six months, and > 2 years post-stroke. DSD-predicted language outcomes were significantly correlated with observed behaviour at the <2 weeks timepoint, but no other significant correlations between DSD-predicted scores were identified. Importantly, DSD-predicted verbal memory and visuospatial domain scores were not significantly correlated with observed behaviour at any of the considered timepoints (minimum p-value = 0.33). Across all tests and timepoints, DSD-predicted scores had an average Mean Absolute Error (MAE) of 0.21 (SD = 0.13, range = 0.04-0.43), with the highest errors occurring between predicted and observed memory scores. Larger stroke lesions were associated with higher MAE, indicating that the DSD performance was modulated by stroke severity.

Overall, these results indicate that the DSD did not yield informative predictions of long-term cognitive outcomes in this external dataset. This finding provides an important illustration of potential overfitting issues within cognitive outcome prediction models, highlighting the need for caution when aiming to predict long-term post-stroke cognitive outcomes and further external validation of proposed models.

## Introduction

Several recent studies have reported on predictive relationships between specific patterns of lesion induced disconnection and long-term cognitive outcomes.^1–3^ This work has informed the development of cognitive outcome prediction tools such as the Disconnection Symptom Discover (DSD).^3^ While the DSD has been found to generate accurate outcomes to a certain extent (approx. 20% variance explained) for selected language scores in an external dataset,^4^ it remains unclear whether this result can be replicated across a wider range of cognitive scores in representative stroke cohorts, and by independent researchers. This study aims to address this gap by conducting an independent, multi-domain evaluation of the DSD’s predictive accuracy throughout the time course of stroke recovery.

Post-stroke cognitive impairment is a common and debilitating consequence of stroke, which is estimated to affect up to 90% of stroke survivors.^5–10^ PSCI is highly heterogenous and can involve independent (or co-occurring) deficits across cognitive domains such as attention, memory, language, and executive function.^11,12^ Stroke survivors with PSCI experience varying degrees of spontaneous recovery^13,14^ with cognitive trajectories differing markedly across different PSCI deficits and individuals.^9,15^ For example, post-stroke visuospatial neglect has a high spontaneous recovery rate with approximately 53% of cases recovering within the first six months post-stroke.^16–19^ However, this group-level result is not consistent across all individuals as recovery rates differ between left vs. right-lateralised neglect, severe vs. mild neglect, and body-centred vs. object-centred neglect.^16,20^ Similar patterns of heterogeneity have been identified across a wide range of different PSCI impairments including language,^21,22^ motor function,^23,24^ and memory.^25^ This variability presents a key challenge for clinical prognostication and recovery planning, as expected individual cognitive outcomes are ultimately unknown.

Previous research has addressed this issue by identifying factors which may help predict differential cognitive outcomes post-stroke. Demographic and clinical characteristics such as age, education, and stroke severity factors are established predictors of long-term cognitive outcomes, though baseline cognition was found to be the strongest predictor.^26^ Indeed, recent efforts for prediction models using baseline cognitive information have shown to provide promising individual level predictions.^27^ Studies have also indicated that brain health measures (e.g. atrophy severity and white matter integrity) are key predictors of cognitive recovery outcomes.^28,29^ In addition, recent work has identified specific patterns of lesion damage acting as granular indicators for domain-specific outcome prediction.^30–33^ While some studies have reported on focal lesion correlates associated with differential cognitive outcomes,^22^ network-level approaches to identify specific patterns of structural and functional disconnection which may help predict cognitive outcomes are gaining strong traction.^1,34–36^ For example, Bowren et al.^1^ conducted a large-scale study reporting that specific disconnection patterns can explain between 6.5-34.4% of variance in 12-month cognitive performance across a variety of language and motor measures. This conclusion has been supported by a range of additional studies identifying disconnection profile as a significant predictor of long-term cognitive outcomes across a wide range of cognitive domains including language, attention, motor skills, and global cognitive outcomes.^2,37–41^

This association between disconnectivity and cognitive outcomes has inspired recent work to explore the extent to which this relationship can be leveraged to support individualised outcome predictions. The Disconnectome Symptom Discoverer (DSD^3^) is an openly available tool which predicts cognitive outcomes based on lesion-derived measures of white matter disconnectivity. To generate predictions, users simply input a probabilistic disonnectome map generated based on a binarised lesion mask.^3^ The DSD then provides individualised predictions of long-term post-stroke performance on 86 different scores from standard neuropsychological measures assessing language, verbal/visuospatial memory, visuospatial attention, motor function, and pain/sickness.^3^ These predictions are generated by machine learning models which were trained on 1-year post-stroke behavioural data from 119 first-time stroke survivors, and was cross validated using three patient samples.^3^ In the original validation study, DSD predictions were found to capture a significant portion of the variance in follow-up cognitive scores (mean absolute error < 20% within 20 held-out patient datasets). Subsequent validation analyses confirmed that DSD predicted semantic fluency scores were significantly correlated with observed behaviour in two additional held-out datasets (R^2^ = 0.18, n = 26, n = 193).^3^ Notably, the DSD is openly available and can be run using data which is routinely collected during stroke care. While the prognostic value of disconnection-based outcome predictions is debated^42–44^, the DSD could potentially provide a promising individualised prediction approach.

A recent independent validation study has indicated that DSD cognitive outcome predictions may not always be consistent with observed scores. Hope et al.^45^ evaluated the DSD’s language outcome prognostic accuracy in an independent sample of 314 stroke survivors from the Predicting Language Outcomes and Recovery After Stroke (PLORAS)^46^ dataset. While correlations between DSD predicted scores and observed language scores obtained at 1-year post-stroke were not significantly different than those reported in the original validation study, these correlations were significantly weaker when DSD predictions were compared to scores collected 5-years post-stroke^4^. This replication study indicates that time post-stroke may be a critical factor modulating DSD accuracy, but it is unclear whether this result is consistent within DSD predictions for non-language cognitive domain. Additionally, Hope et al.^45^ did not assess model accuracy in terms of prediction error which is also critical to understanding model accuracy.^47^

Additionally, it is not yet clear whether the results of previous DSD validation analyses are replicable in more representative stroke samples. For example, the average age of DSD sample cohorts has been between 53 and 64,^3,4^ while the average stroke age in most countries is >70. To our understanding, DSD validation studies have also excluded patients who are unable to complete extensive neuropsychological testing and thereby are likely biased towards minor stroke as the lengthy and complex testing is likely to have excluded more cognitively impaired stroke survivors.^48,49^ To be considered clinically useful tools, prognostic indicators must be able to produce accurate impairment predictions within representative patient cohorts,^42^ particularly to help predict recovery in those with clear acute cognitive impairments. It therefore remains critically important to investigate whether the DSD can support meaningful multi-domain cognitive outcome predictions in a clinically representative independent cohort.

The present study addresses these key research gaps by providing an independent, external, multi-domain validation of DSD predicted cognitive outcomes. We used the DSD to generate predicted cognitive outcomes for a representative stroke cohort (n = 74) and evaluated the accuracy of these predictions across three cognitive domains (language, memory, and visuospatial attention) assessed at three distinct time points post-stroke (<2 weeks, 6 months, >2 years). We aimed to provide a rigorous, independent and out-of-sample validation test of the DSD, providing novel insight into this promising tool’s potential prognostic utility in real-world clinical stroke settings.

## Materials and Methods

### Participant Sample

This study is a retrospective analysis of stroke survivors recruited in the OX-Chronic study from the John Radcliffe Hospital (UK, 2012-2020).^50,51^ All participants provided informed consent in line with the Declaration of Helsinki (REC references:14/LO/0648, 18/SC/0550, 12/WM/00335). The OX-Chronic study included all stroke patients who could sustain attention for at least 20 minutes and had a good understanding of English. From this dataset, individuals with available routinely collected neuroimaging (CT scans) demonstrating visible lesions were included in the present study (mean stroke-scan interval = 2.27 days (SD = 6.77)). Patients with imaging which could not be accurately normalised were excluded.

Overall, 74 participants (mean age = 69.18 (SD = 12.25), 59.5% male, 89.2% right-handed) were included in the present study. This sample included both participants with first-time stroke (63.5%) and recurrent strokes (36.5%). Stroke types were reported as 75.7% ischemic and 24.3% haemorrhagic. Stroke locations were reported as 35 left, 34 right, and 5 bilateral. Cognitive assessment was conducted during acute hospitalisation (<2 weeks post-stroke) (M = 4.81 days post-stroke (SD = 5.95)), at approximately six months post-stroke (M = 204.28 days, SD = 38.62), and at >2 years post-stroke (M = 4.56 years, SD = 2.19, range = 2.0 – 9.4 years). A priori power calculations indicated that a minimum sample of 49 was needed to detect the smallest reported relevant effect size (r = 0.35) reported by Tazzol et al.^3^ in a one-tailed correlation test (alpha level = 0.05). Lesion distribution and sample-level disconnection profile is summarised in Figure 1.

**Figure 1:**
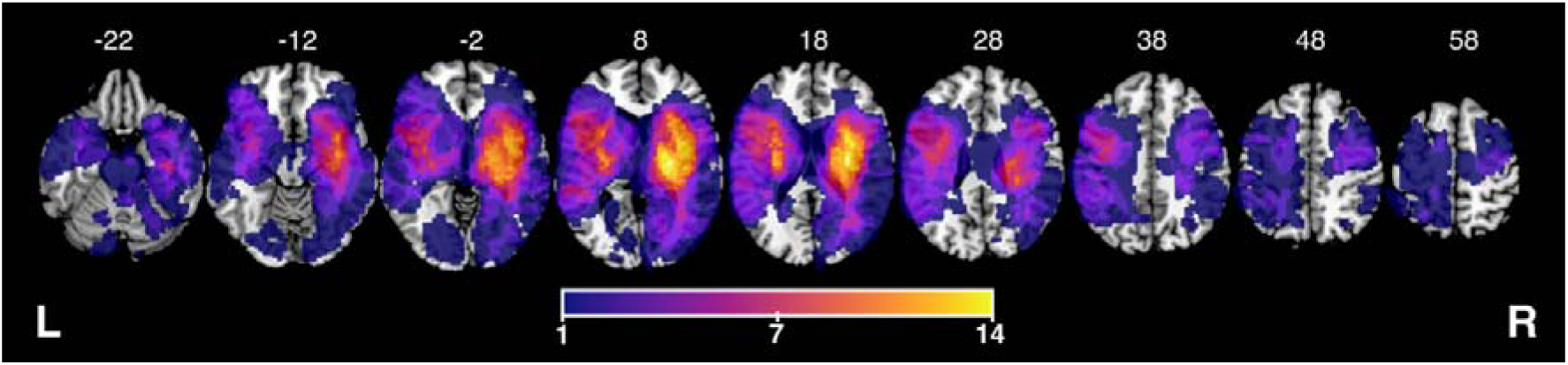
Lesion overlay for the included sample (n = 74). Colour represents the number of patients with lesions impacting each area. Axial slices between MNI X -22-58 are presented.

Table 1 presents detailed stroke characteristics and demographics for this study’s sample in comparison to the sample originally reported by Talozzi et al.^3^ To evaluate sample comparability continuous variables were compared using independent t-tests and categorical variables were compared using chi-squared tests. The present study’s cohort was not significantly different from the cohort reported by Talozzi et al.^3^ in terms of sex, handedness, education level, and lesion laterality (p-value range = 0.54-0.98). However, this study’s sample was significantly older (69.18 vs 54; *t*(142) = 8.70, *p* < .001) and had imaging data which was collected significantly earlier after stroke (2.27 vs 14 days; *t*(174) = -10.90, *p* <.001).

**Table 1:** Demographic and stroke characteristics. Study sample in comparison to the original sample from Talozzi et al. ^3^ Continuous data are reported in terms of means and standard deviations, while categorical variables are reported as percentages.

| Characteristic | OX-Chronic | Washington |
| --- | --- | --- |
| *n* | 74 | 119 |
| Demographic characteristics |  |  |
| Sex | 59.5% male / 39.2% female / 1.4% unknown | 54.6% male / 45.4% female |
| Age, years | 69.18 ± 12.25 (30.35 - 92.21) | 54 ± 11 (19 - 83) |
| Education, years | 13.36 ± 4.03 (9 - 28) | 13.2 ± 2.5 (5 - 20) |
| Handedness | 89.2% right-handed / 9.5% left-handed / 1.4% ambidextrous | 91.6% right-handed / 8.4% left-handed |
| Stroke characteristics |  |  |
| Stroke type | 75.7% ischemic / 24.3% hemorrhagic | - |
| Lesion hemisphere | 45.9% right / 47.3% left / 6.8% bilateral | 46% right / 54% left |
| Lesion volume, cm<sup>3</sup> | 38.42 ± 31.43 | 34 |
| Neuropsychology assessment relative to stroke | <2 weeks: 4.81 days ± 5.95<br>Six months: 204.28 days ± 38.62<br>>2 years: 4.56 years ± 2.19 | 393 days ± 56 |
| Neuroimaging relative to stroke, days | 2.27 ± 6.77 | 14 ± 8 |

#### Neuroimaging Data

First, predicted behavioural scores were generated by the DSD based on disconnectome maps. To generate these maps, stroke lesions were manually delineated on native space CT scans in line with the standard procedure reported by Moore.^52^ CT is the international standard neuroimaging method for acute stroke^53–55^ and is recommended for use in lesion mapping analyses.^56,57^ Critically, CT-derived lesion masks have been demonstrated to be non-inferior to MRI-derived lesion masks in lesion mapping,^52^ and have been shown to improve lesion-mapping power and sensitivity by improving lesion coverage over analyses which use MR alone.^58^ For haemorrhagic strokes, delineations included both the intracerebral blood and surrounding peri-focal hypodense tissue. The resulting lesion masks were smoothed in the z-direction at 5 mm full-width at half maximum to reduce noise, and then binarised using a threshold of 0.5. These masks were reoriented to standard anatomical orientation and normalised into MNI152^59^ 2x2x2 mm stereotaxic space, as required by the DSD pipeline. This procedure used Statistical Parametric Mapping^60^ and Clinical Toolbox functions.^61^ Final lesion masks were all visually inspected for quality prior to inclusion. Data which was unable to be accurately normalised was excluded from this study.

In line with standard procedure for using the DSD toolkit, normalised lesion masks were used to generate disconnectome maps using tools from the Brain Connectivity and Behaviour (BCB) Toolkit.^62^ This process compares each lesion mask to 7T MRI diffusion-weighted images from 178 healthy controls to produce a whole-brain map where each 2x2x2 mm voxel encodes the probability of an underlying white matter connection being disrupted due to the lesion (see Figure 2B). The resulting disconnectome maps were then input into the DSD model to generate predicted scores for cognitive outcomes. Full details of the DSD model pipeline are described in Talozzi et al.^3^

**Figure 2:**
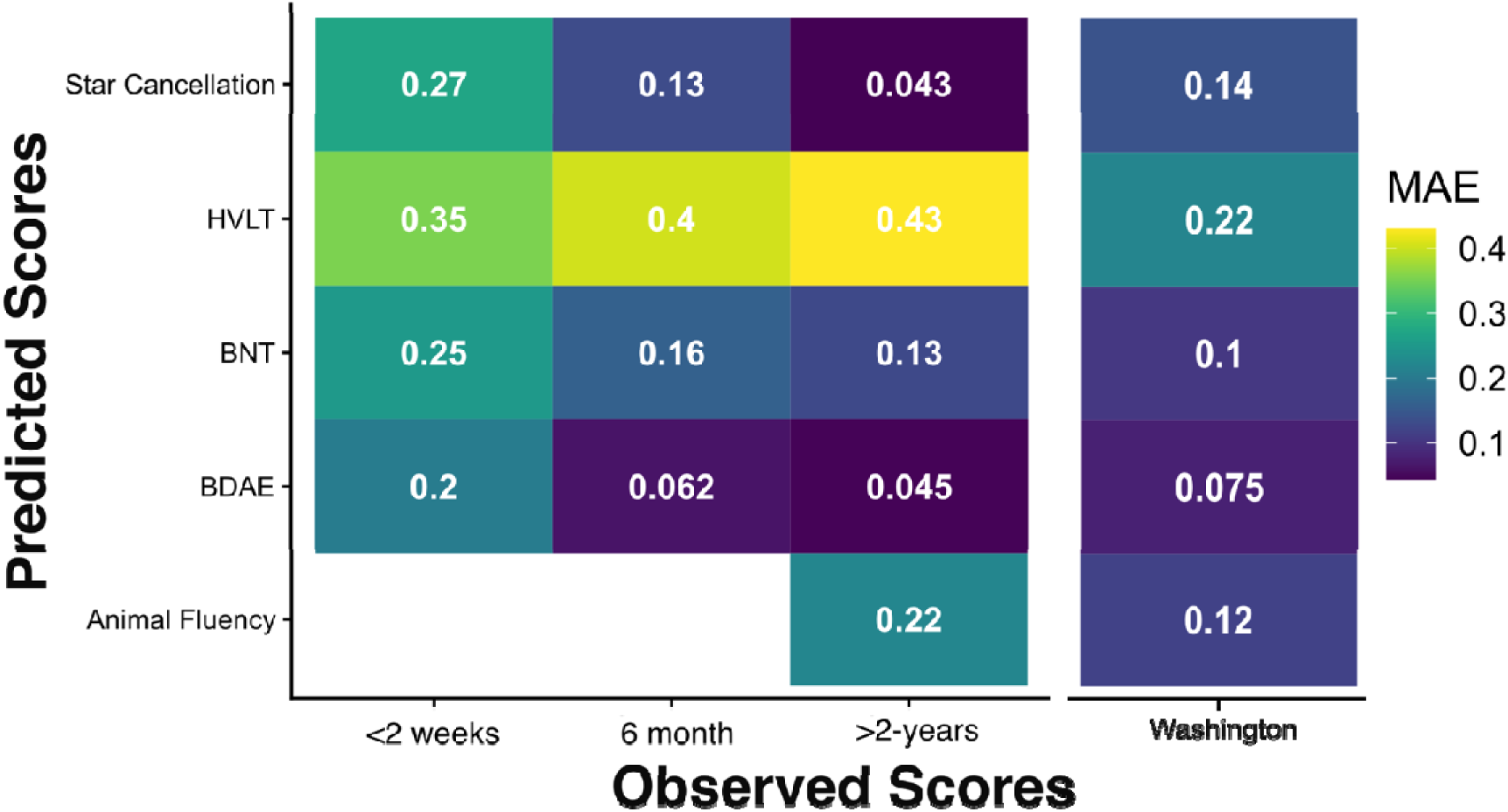
Mean absolute error (MAE) for predicted versus observed scores across all tests and timepoints. Data from the present cohort (<2 weeks, 6 months, and >2 years columns) is reported alongside results in the original validation study (Washington column) for reference.

#### Actual and Predicted Cognitive Outcome Data

Predicted cognitive outcomes were compared to observed cognitive data collected from the patient sample across three cognitive domains: visuospatial attention, verbal memory, and language. Where possible, predicted scores were exactly matched to identical collected neuropsychological tests (Table 1). In other cases, predictions were compared to performance on analogous (but not identical) neuropsychological tests (Table 2) in line with previous DSD validation approaches.^45^ These analogous tests showed high convergent validity with the same underlying cognitive functions as the predicted test scores.^48^ Though differences in test materials and difficulty mean that analogous scores should not be expected to be identical to their paired predicted scores, these measures are expected to strongly correlate.

**Table 2.** Predicted and Observed Neuropsychological Test Scores. DSD predicted outcomes and paired observed neuropsychological test at each timepoint.

| Predicted test | Observed test |  |  |
| --- | --- | --- | --- |
|  | <2 weeks | Six months | >2 years |
| **Visuospatial ability** |  |  |  |
| Star Cancellation Test | OCS Broken Hearts | OCS Broken Hearts | Star Cancellation Test |
| **Verbal memory** |  |  |  |
| HVLT | OCS Verbal Recall | OCS Verbal Recall | OCS Verbal Recall |
| **Language** |  |  |  |
| BNT | OCS Picture Naming | OCS Picture Naming | BNT |
| BDAE Sentence Reading | OCS Sentence Reading | OCS Sentence Reading | OCS Sentence Reading |
| Animal Fluency Test | - | - | Animal Fluency Test |

This study included observed behavioural data from the Oxford Cognitive Screen (OCS; Demeyere et al., 2015), Birmingham Cognitive Screen (BCoS),^63^ Behavioural Inattention Test Star Cancellation Test (BIT),^64^ Boston Naming Test (shortened version; BNT; Mack et al., 1992), and the Animal Fluency Test.^66^ The OCS was completed during acute testing <2 weeks post stroke, the BCoS or OCS was completed during testing at six months (17 OCS, 57 BCoS), and testing at the >2 years timepoint included the OCS, BIT Star Cancellation Test, BNT (shortened version), and Animal Fluency Tests. Where the BCoS was used, BCoS scores from subtests analogous to the relevant OCS subtests were used in analysis. Data from the OCS Cancellation (Broken Hearts Test), Verbal Recall, Picture Naming, and Sentence Reading subtests was considered in this study. Detailed descriptions of these OCS subtests and scoring procedures are reported in detail elsewhere.^48,67^ Task details and scoring instructions are provided in Supplementary Materials.

See Table 1 for a summary of which observed and predicted tests are compared at each assessment timepoint.

### Statistical Analysis

Raw scores for both observed and predicted data were converted into proportions of the maximum possible score for each test to standardise scaling across test versions and facilitate comparability. Descriptive statistics (M ± SD) for the scaled observed and predicted scores were calculated for each test and timepoint. In cases where individuals were missing data for specific tests, these patients were excluded from relevant statistical analyses (see Table S1). Predicted score accuracy was evaluate using Mean Absolute Error (MAE). These MAE values were compared to corresponding results reported in the original DSD Washington dataset.^3^

To assess the predictive ability of the DSD, predicted scores were correlated with observed scores at each time point. The resulting correlation coefficients (*r* values) were qualitatively compared to analogous measures reported in the DSD Washington dataset^3^ and validation PLORAS dataset.^45^ In line with standard interpretation guidelines,^68^ *r* values > 0.7 were considered to represent strong correlations, *r* values between 0.3-0.7 were considered moderate, and *r* values < 0.3 were considered weak correlations.

To determine whether resulting correlations were statistically comparable to those in the original study, Fisher’s r-to-z transformations were performed, comparing the generated correlation coefficients with those reported from the Washington data. This method enables direct statistical comparison between studies and is consistent with previous DSD validation work (Hope et al., 2024). To evaluate the extent to which prediction accuracy may be impacted by individual differences, average MAE across all subtests and testing times (per-participant) was correlated with age and lesion volume. A t-test was also conducted to compare average MAE between patients with first-time vs. recurrent stroke diagnoses. A significance threshold of *p* ≤ .05 was applied to all statistical tests. All analyses were conducted in R version 2024.12.0.

## Results

Descriptive statistics for the observed and predicted scores at each time point are reported in Table 3. MAE for predicted vs. observed scores at each time point are reported in Figure 2 and Table S1. Across all tests and timepoints, the average MAE was found to be 0.21 (SD = 0.13, range = 0.043 – 0.43). MAE was largest for the HVLT at all timepoints, ranging between 31.0% to 35.0% error, compared to 21.8% in the Washington dataset. For Star Cancellation Test, BNT, and BDAE Sentence Reading, MAE was highest at the <2 weeks timepoint (20.5% - 30.1%) and lowest at the chronic timepoint (4.2% - 9.5%). The MAE reported in the Washington dataset varied between 7.5% and 14.0% for these tests. Across all considered tests and timepoints MAE was higher in this cohort than in the original Washington University cohort in 9/13 cases.

**Table 3:** Observed and Predicted Cognitive Outcome Scores. Means and standard deviations for all observed and predicted cognitive outcomes across considered timepoints. Values are expressed in terms of proportion correct, with the exception of the Animal Fluency Test which is reported in terms of raw values.

| Test | <2 weeks | Six months | 2> years | Predicted |
| --- | --- | --- | --- | --- |

**Table 3: Observed and Predicted Cognitive Outcome Scores.**
|  |  |  |  |  |
| --- | --- | --- | --- | --- |
| Star Cancellation Test | $0.69 \pm 0.31$ | $0.89 \pm 0.13$ | $0.96 \pm 0.12$ | $0.99 \pm 0.00$ |
| **Verbal Memory** |  |  |  |  |
| HVLT | $0.77 \pm 0.31$ | $0.92 \pm 0.17$ | $0.91 \pm 0.20$ | $0.63 \pm 0.15$ |
| **Language** |  |  |  |  |
| BNT | $0.73 \pm 0.33$ | $0.89 \pm 0.19$ | $0.93 \pm 0.13$ | $0.91 \pm 0.04$ |
| BDAE Sentence Reading | $0.79 \pm 0.35$ | $0.95 \pm 0.20$ | $0.97 \pm 0.12$ | $0.95 \pm 0.03$ |
| Animal Fluency Test | - | - | $15.64 \pm 6.28$ | $18.05 \pm 3.26$ |

Correlations between predicted and observed scores are reported in Figure 3. At the <2 weeks timepoint, DSD predicted language scores (BDAE and BNT) were significantly moderately correlated with the matched observed scores. Correlations between predicted verbal memory and visuospatial attention scores were not significant at the <2 weeks stage. At the six months and >2 years timepoints, no significant associations were observed between predicted and observed scores (minimum p-value = 0.07), with correlation coefficients ranging between .21 and -0.8.

**Figure 3:**
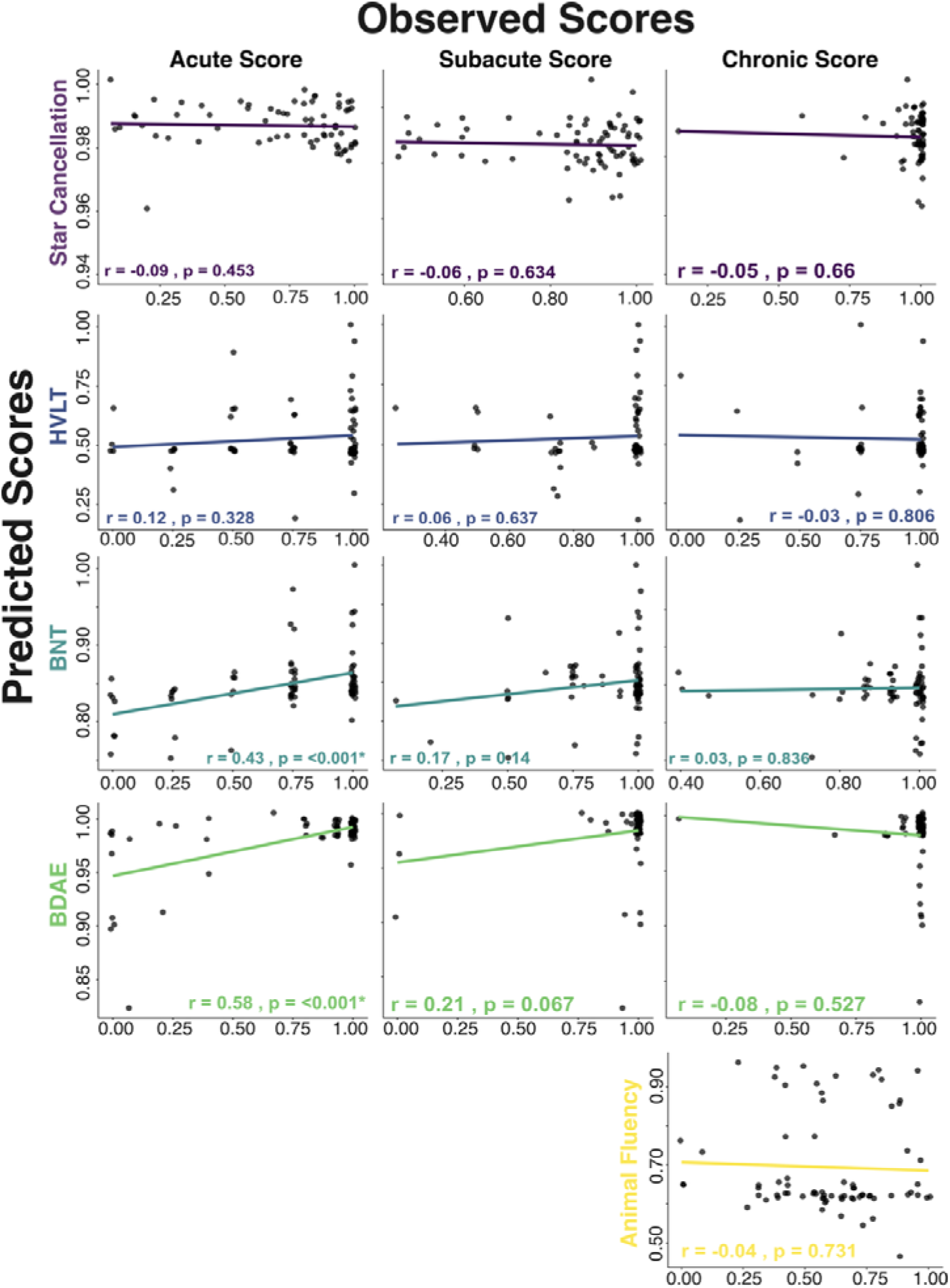
Scatter plots depicting correlations between observed and predicted scores for all tests across different timepoints. Observed and predicted scores are reported as a proportion of the maximum possible score. Dots denote individual participant scores and lines summarise correlation results. Correlation statistics (r and p-value) are reported in the lower portion of each panel. Significant correlations (p < 0.05) are denoted by stars.

For all tests at the chronic timepoint, Fisher’s r-z transformation demonstrated that correlations between observed and predicted scores in the present cohort have significantly lower correlation values than in the Washington dataset (Table 4). At the six months timepoint, only the Star Cancellation Test and HVLT showed lower correlations, whereas the BNT and BDAE Sentence Reading task did not perform significantly differently from the Washington dataset. At the <2 weeks timepoint, the Star Cancellation Test showed a lower correlation value relative to the Washington cohort, while the HVLT, BNT, and BDAE Sentence Reading task did not significantly differ.

**Table 4.**
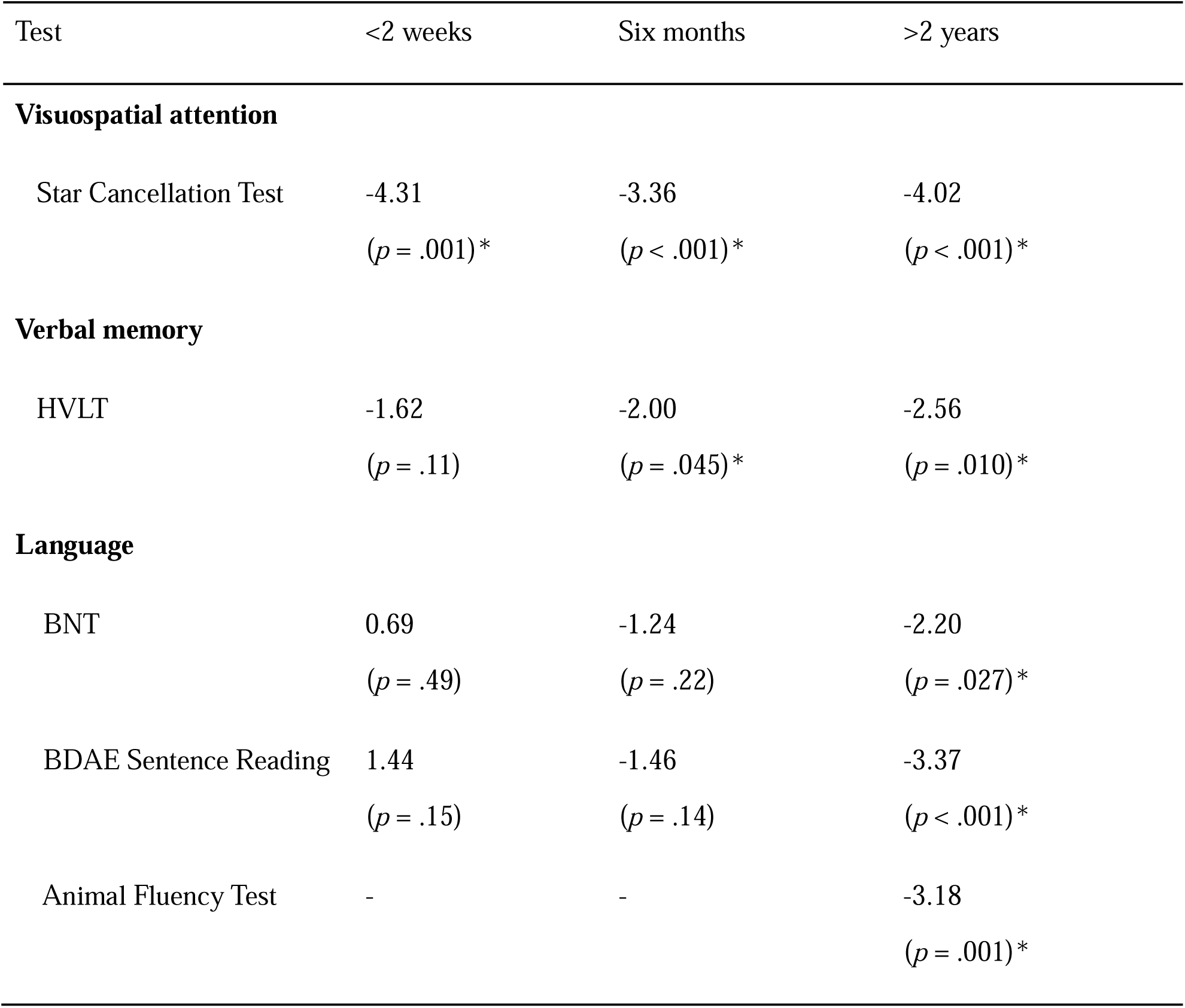
Correlation r-value comparisons between this study’s cohort and the original Washington Sample ^3^. Fisher’s r-to-z transformation scores are reported alongside p-values. Significant differences (denoted by stars) highlight cases in which correlations significantly differ between cohorts.

Finally, analyses were conducted to explore the extent to which individual differences may modulate DSD prediction accuracy. Average per-participant MAE was not significantly correlated with age (Figure 4). However, this value was found to be significantly, positively correlated with log lesion volume indicating that DSD prediction accuracy decreases in patients with larger, more severe strokes (Figure 4). Average MAE was also not significantly different between participants with first-time stroke and participants with recurrent stroke *(t*(50.7) = -0.17, *p* = 0.859, 95% CI: -0.037 - 0.031).

**Figure 4:**
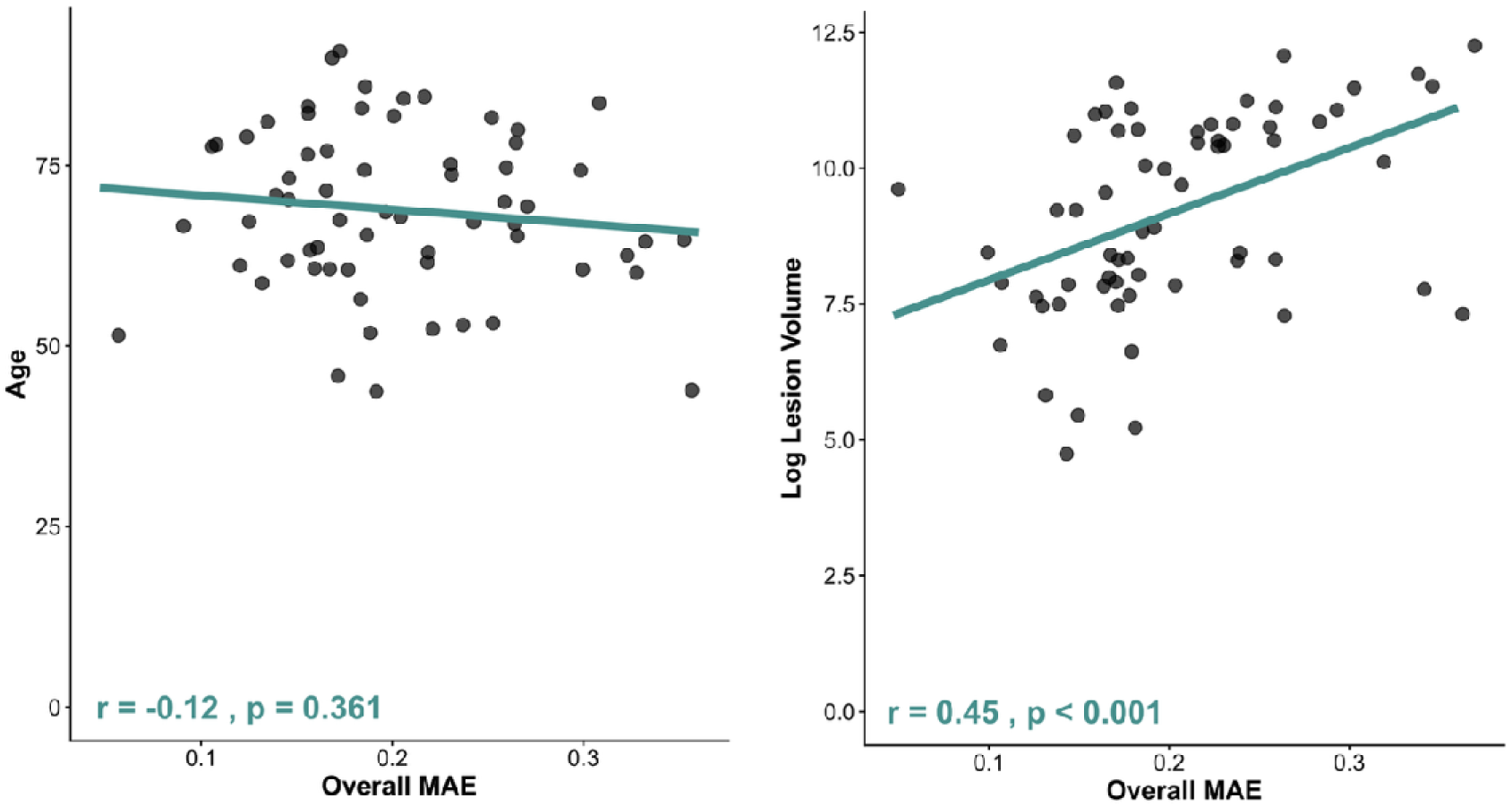
Relationship between age, lesion volume, and DSD MAE. Observed and predicted scores are reported as a proportion of the maximum possible score. Dots denote individual participant scores and lines summarise correlation results. Correlation statistics (r and p-value) are reported in the lower portion of each panel. Significant correlations (p < 0.05) are denoted by stars.

## Discussion

This study presents an independent, external, multi-domain validation of DSD predicted cognitive outcomes across three distinct timepoints in stroke recovery. In line with previously reported validation results, DSD-predicted language outcomes were significantly correlated with observed behaviour.^3,4^ However, this relationship was only present when observed behaviour was collected at the most acute timepoint (<2 weeks post-stroke). Additionally, predicted verbal memory and visuospatial domain scores were not significantly correlated with observed behaviour at any of the considered timepoints. Overall, these results indicate that the DSD did not yield informative predictions of long-term cognitive outcomes in this external dataset. This result highlights relevant limitations in the DSD’s predictive capabilities which have important implications for future research and clinical applications of this model.

### The DSD may not yield informative cognitive outcome predictions

This study’s analyses of DSD predicted language outcomes are consistent with previous external validation work. Hope et al.^28^ found the strength of correlation between DSD-predicted outcomes and observed behaviour weakened as the interval between stroke and behavioural testing increased. This is supported by the present study, as correlations between observed and predicted language scores were only significantly present at the earliest post-stroke timepoint (<2-weeks post-stroke) and were insignificant at later timepoints. There are several potential explanations for this time-dependent decline in predictive power. Brain-behaviour relationships are strongest when both imaging and behavioural data are collected at acute timepoints.^56,69,70^ The impact of recovery processes such as functional reorganisation increases as time post-stroke increases, introducing noise into relationships between focal lesion damage patterns and behavioural scores as time progresses.^56,69^ Chronic outcomes are also influenced by a broad range of factors outside of lesion location, including lifestyle, psychological status, and other health events.^71–73^ The identified significant correlation between DSD predicted language scores and observed acute behaviour provides a promising sanity-check indicating that underlying metrics of white matter connectivity associate with language. However, the prognostic utility of this relationship is likely limited as the goal of predictive modelling is to predict future outcomes, not to report on current impairment levels.^74^

Importantly, DSD predictions of visuospatial attention, verbal memory, and post-acute language scores were not found to be correlated with observed behaviour. In terms of predicted visuospatial attention scores, this replicates the results of the original DSD validation study which reported non-significant relationships between predicted and observed Star Cancellation Test scores.^3^ In this sample, DSD predicted Star Cancellation Test total scores exhibited low variability and were uniformly high with 94.6% of patients predicted to miss less than one target. This is likely due to a ceiling effect, present in the long term data due to the strong recovery profiles for visuospatial neglect,^17^ thus limiting the strength of possible correlations with observed data.

This study represents the first independent validation analysis of DSD-predicted memory scores. However, DSD predicted verbal memory scores were not significantly correlated with observed behaviour at any of the considered timepoints. Importantly, past research investigating the recovery trajectory of post-stroke memory impairments has identified several factors outside of lesion location which may play a key role in determining memory outcomes. For example, Hobden et al.^29^ found that global cortical atrophy severity was strongly associated with poor post-stroke memory recovery outcomes. Recent work has also suggested that many cases of memory impairment which are detected in post-stroke cognitive screening may be unrelated to the acute stroke event.^75,76^ This is because many patients may experience undetected cognitive decline prior to the stroke event which may introduce noise into lesion-behaviour analyses.^77^ This effect may help account for why verbal memory predictions were poor even compared to acute observed behaviour.

In addition to correlation measures of DSD performance, it is also important to interpret model performance in terms of absolute accuracy of scores. Correlation analyses report on whether model predictions capture directional and relative rank relationships, for predicted scores to be clinically useful, these values must accurately reflect the absolute level of impairment in individuals. In this study, predictive correlations were highest at the acute stage, yet the MAE was also greatest at this timepoint indicating poor precision for individual-level estimates. Lower MAE values at six months and >2 years post-stroke likely reflect closer alignment to the original one-year post-stroke outcomes on which the DSD model was trained,^3^ given that recovery trajectories tend to be more stable across these timepoints.^13,14^

### Factors modulating DSD predictive accuracy

Overall, the DSD-predicted cognitive outcomes were not correlated with the observed long-term cognitive outcomes post-stroke. This finding diverges from both the original DSD validation study and a previous DSD external validation study on selected language outcomes.^45^ It is possible that a portion of this divergence may be related to cohort differences between the present study and this previous work. Mainly, this study’s cohort was, on average, 15 years older at stroke onset compared to the sample used to train and test the DSD. Age at stroke onset is a well-established predictor of poor cognitive recovery after stroke as well as lower baseline cognitive ability.^10,33,78^ The age distribution in the present sample is more representative of the stroke population, as average age of stroke is generally between 68 and 74.^79^ It is plausible that the DSD may generate more accurate outcome predictions for younger relative to older stroke survivors. However, DSD accuracy was not significantly correlated with stroke age in our sample.

Additionally, the present sample included some individuals with recurrent stroke, whereas some previous DSD validation work has only included first time stroke survivors.^3^ Recurrent strokes tend to result in greater cumulative neuronal damage and are associated with more severe cognitive impairment than first strokes,^80,81^ and this could plausible lead to greater variability of the observed cognitive impairments and lesion masks. However, no significant difference in model error was present between participants with first vs. recurrent strokes in this sample. Notably, lesion volume was significantly correlated with DSD accuracy with patients with smaller lesions receiving more accurate DSD outcome predictions. This study’s average lesion volume (38 cm^3^) is only slightly larger than that in the sample used to train the DSD (34 cm^3^), but the lesion volume for all external validation cohorts has not been reported (Hope et al., 2023). Studies tracking long-term cognitive outcomes often report on comparatively mild stroke samples, partly because neuropsychological testing burden excludes more severe stroke, and partly as more severe patients are likely to have higher attrition rates over time.^82^ The results of this study indicate that lesion volume may be a key factor modulating the DSD’s prognostic accuracy.

There are also several design differences between this project and previous DSD validation work which may help to account for diverging results. The present study considered long-term cognitive outcomes collected at two weeks, six months, and two years post-stroke while the DSD was trained to predict cognitive outcomes at one-year post-stroke. For many cognitive domains, the bulk of cognitive recovery has been found to occur within the first six months to 1 year following stroke, with cognitive performance at six months, one year, and two years post-stroke generally being highly correlated.^18,77,82,83^ This implies that differences in testing time alone are not sufficient to account for the failure to replicate DSD validation results. Differences in neuropsychological tests analysed may also have affected comparability between observed and predicted scores in this study. Previous DSD external validations have provided evidence that DSD predictions may be extended to analogous, but not identical neuropsychological tests scores.^45^ However, the OCS is a brief cognitive screen containing simple screening tasks which yield a restricted range of possible scores relative to extended neuropsychological batteries.^48^ For example, HVLT verbal recall (predicted by the DSD) requires participants to remember 12 items for approximately 20 minutes,^84^ while the OCS verbal recall task requires participants to recall four words from a sentence for approximately 5 minutes. This difference in score ranges and difficulty could help explain differences between observed and predicted scores. However, DSD predicted scores were also not found to be correlated with observed behaviour for chronic star cancellation and animal fluency scores, both of which are the exact same tests as used in the original DSD modelling.

Additionally, the DSD was trained and validated using lesion masks derived from MRI imaging, but this study used CT-derived lesion masks to generate outcome predictions. CT and MR-derived lesion masks have been shown to perform comparably in lesion mapping analyses,^85,86^ but residual differences in delineation precision and normalisation may introduced noise into DSD predictions.^61,87,88^ Finally, the interval between imaging and initial behavioural testing was shorter in the present study relative to the original DSD sample (2 days vs. 14 days respectively). The delayed imaging in the DSD training dataset may have potentially captured brain lesions further along in healing process and therefore be trained to a different neuroanatomical profile than what is observed in earlier neuroimaging. Each of these timing and neuroimaging factors are important to consider in the context of the DSD’s potential clinical applications as neuroimaging timing is subject to variation in general stroke practice.^89^ Cognitive outcome predictions which are dependent on the precise timing and modality of neuroimaging may not be as useful as a more generalisable prediction approach.

While the results of this study illustrate key limitations in the DSD’s predictive accuracy, several important questions remain open to future research. First, this study evaluated a small subset of the 86 neuropsychological test scores predicted by the DSD. Future research can aim to conduct more large-scale validation projects to investigate whether the results of this study generalise to a wider set of DSD predicted outcomes. Second, the results of this study indicate that individual differences (such as lesion size) may plan a key role in modulating the DSD predictive accuracy. This study’s sample size is not sufficient to support detailed patient subgroup analyses, but future work can aim to conduct more detailed explorations of how differences such as lesion size, premorbid brain health, and lesion location may ultimately impact DSD accuracy. Future development work can aim to explore whether retraining the DSD on larger and more clinically representative datasets, incorporating recurrent strokes, older participants, and earlier imaging. Additionally, future work may want to integrate the DSD model with established demographic, behavioural and clinical characteristics to improve predictive capacity.^26,42^ For complex neuroimaging-based biomarkers of cognitive outcome to have prognostic value in real-world clinical environments, it is important for these measures to provide information which is not already captured by more readily available, established prognostic indicators. Future research can aim to address this key gap by directly comparing the value of DSD-derived outcome predictions to basic established predictors of cognitive recovery outcome such as such as age, stroke severity, acute cognitive profile, and premorbid health.^26,27,90^

Overall, this study highlights key limitations in the DSD’s ability to accurately predict long-term cognitive outcomes. While some degree of this reduced accuracy may be related to demographic, clinical, and neuropsychological testing difference between this cohort and previously reported samples, these factors alone are unlikely to fully account for the consistent lack of correlation between DSD predicted outcomes and observed behaviour documented here. Future research can aim to provide more detailed insights into the factors modulating DSD accuracy to identify avenues to improve outcome prediction accuracy. Clinically useful prognostic measures must provide reliable and accurate outcome predictions in representative cohorts. In this context, it is important for researchers and clinicians to exercise extreme caution when using models such as the DSD to estimate future levels of cognitive impairment in individual stroke survivors.

## Data Availability Statement

All data and code associated with this project is openly available on the Open Science Framework (https://osf.io/psbue/).

## Acknowledgements

This project was supported by the Oxford Cognitive Screening Programme. Specifically, we would like to acknowledge Dr. Georgina Hobden for her contributions to generating the cortical atrophy rating used in this study.

## Author Contributions

In line with the CRediT taxonomy, LK was responsible for Data Curation, Fromal Analysis, Investigation, Methodology, Visualisation, and Writing – original draft. MJM was responsible for Conceptualisation, Data Curation, Investigation, Methodology, Supervision, Visualisation, and Writing – reviewing & editing. ND was responsible for Conceptualisation, Resources, Supervision, and Writing – reviewing & editing

## Abbreviations

Disconnection Symptom Discoverer: (DSD)
Mean Absolute Error: (MAE)
Oxford Cognitive Screen: (OCS)
Boston Diagnostic Aphasia Examination: (BDAE)
Hopkins Verbal Learning Task: (HVLT)
Boston Naming Task: (BNT)

