## Supplemental materials for "An independent, multi-timepoint evaluation of Disconnection Symptom Discoverer cognitive outcome prediction accuracy in stroke"

### **Supplementary Materials**

**Neuropsychological Testing Details:**

The OCS Broken Hearts Test is a test of visuospatial attention in which participants are presented with a search array of complete hearts (targets) and incomplete hearts (distractor) and are instructed to cross off all targets (maximum score = 50). The total number of target misses in the OCS Broken Hearts Task was compared to the DSD’s prediction of total target misses in the BIT Star Cancellation Test. The BIT Star Cancellation (Wilson et al., 1987) assesses visuospatial attention by asking participants to select the small stars across an A4 sheet of paper or screen, while ignoring large stars or letters (maximum score = 54). For chronic timepoint analyses, this predicted score is compared directly to the actual BIT Star Cancellation tests completed by patients.

Three language domain cognitive outcomes were evaluated: sentence reading, picture naming, and verbal fluency. In the OCS Sentence Reading Task, participants are instructed to read a presented sentence out loud (maximum score = 15). Data from this test was compared to the DSD’s predicted score for the BDAE Sentence Reading Test. The BDAE Sentence Reading Test^90^ requires participants to read out 10 sentences of increasing length and complexity. If the whole sentence is read correctly, then it is scored as correct yielding a maximum total score of 10.

In the OCS Picture Naming Test, participants are asked to name common objects/animals depicted in drawings (maximum score = 4). This data is compared to the predicted score for the shortened BNT, which requires participants to name 15 objects from line drawings (semantic cues provided after 20 seconds, maximum score = 15). At the chronic timepoint, patients completed the shortened BNT and this total is directly compared to DSD’s predicted score. The Animal Fluency Test^65^ requires participants to name as many animals as possible in one minute. Participants completed this test at the chronic timepoint, and actual scores are compared to DSD predicted scores.

The OCS Verbal Recall task (a verbal memory assessment) includes four multiple choice questions where participants are asked to select words which appeared in the text read during the earlier sentence reading task (approximately 5-minute delay, maximum score = 4). This observed data was compared to the predicted HVLT score. The HVLT^83^ assesses verbal memory by asking participants to memorise a list of 12 items read aloud. Delayed recall of these words is scored 20–25 minutes (maximum score = 12).

| Test | <2 weeks | | 6 months | >2 years |
| --- | --- | --- | --- | --- |
| **Visuospatial attention** |  |  | |  |
| Star Cancellation Test | 73 | 74 | | 70 |
| **Verbal memory** |  |  | |  |
| HVLT | 73 | 72 | | 71 |
| **Language** |  |  | |  |
| BNT | 74 | 74 | | 71 |
| BDAE Sentence Reading | 73 | 74 | | 71 |
| Animal Fluency Test | - | - | | 74 |

***Table S1.*** *Sample size analysed for each neuropsychological test and timepoint.*

| Test | <2 weeks | 6 months | >2 years | Washington |
| --- | --- | --- | --- | --- |
| **Visuospatial attention** |  |  |  |  |
| Star Cancellation Test | 30.1% | 10.3% | 4.2% | 14.0% |
| **Verbal memory** |  |  |  |  |
| HVLT | 31.4% | 32.9% | 35.0% | 21.8% |
| **Language** |  |  |  |  |
| BNT | 25.7% | 13.8% | 9.5% | 10.2% |
| BDAE Sentence  Reading | 20.5% | 8.3% | 6.4% | 7.5% |
| Animal Fluency Test | - | - | 5.2% | 11.6% |

***Table S2.*** *Mean Absolute Error of Predicted Scores Compared to Observed Scores from the OX-Chronic and Washington Samples.*
